# Early, intensive rehabilitation improves gross motor function after perinatal stroke: results of a randomized controlled trial

**DOI:** 10.1101/2021.07.21.21260801

**Authors:** Caitlin Hurd, Donna Livingstone, Kelly Brunton, Allison Smith, Monica Gorassini, Man-Joe Watt, John Andersen, Adam Kirton, Jaynie F. Yang

## Abstract

**Background:** Perinatal stroke injures motor regions of the brain, compromising movement for life. Early, intensive, active interventions for the upper extremity are efficacious, but interventions for the lower extremity (LE) remain infrequent and understudied.

**Objective:** To determine the efficacy of ELEVATE – Engaging the Lower Extremity Via Active Therapy Early - on gross motor function, as compared to usual care.

**Methods:** We conducted a single-blind, two-arm, randomized controlled trial (RCT), with the Immediate Group receiving the intervention while the Delay Group served as a three-month waitlist-control. A separate cohort living beyond commuting distance was trained by their parents with guidance from physical therapists. Participants were 8 months to 3 years old, with MRI-confirmed perinatal ischemic stroke and early signs of hemiparesis. The intervention was play-based, focused on weight-bearing, balance and walking for 1 hour/day, 4 days/week for 12 weeks. The primary outcome was the Gross Motor Function Measure-66 (GMFM-66). Secondary outcomes included steps and gait analyses. Final follow-up occurred at age four.

**Results:** Thirty-four children participated (25 RCT, 9 Parent-trained). The improvement in GMFM-66 over 12 weeks was greater for the Immediate than the Delay Group (average change 3.4 units higher) and greater in younger children. Average step counts reached 1370–3750 steps/session in the last week of training for all children. Parent-trained children also improved but with greater variability.

**Conclusions:** Early, activity-intensive LE therapy for young children with perinatal stroke is feasible and improves gross motor function in the short term. Longer term improvement may require additional bouts of intervention.

**Clinical trial registration:** This study was registered at ClinicalTrials.gov (NCT01773369).

## Introduction

Perinatal stroke is a cerebrovascular event that occurs between gestational age of 20 weeks and 28 days postnatal, and has an incidence between one in 1600 to 2300 live births^1,2^. It is the leading cause of hemiparetic cerebral palsy (CP), which may involve weakness, spasticity and impaired coordination in the upper and lower extremity on the affected side of the body^3^. The life-long gross motor impairments contribute to long-term musculoskeletal complications, impaired gait, reduced physical activity and participation^4,5^.

Intensive, active approaches to rehabilitation have been effective for improving upper extremity function (e.g. constraint-induced movement therapy [CIMT] and bimanual training, reviewed in ^6–8^). In contrast, active treatment approaches for the lower extremity are limited for young children with CP^7^, although some studies combining upper and lower extremity training show promise^9,10^ or are underway^11^. Optimizing lower extremity function is especially important now, because of the recent reduction in severity of CP among developed countries, resulting in more children with the potential to walk^12^.

Current clinical practice for lower extremity function in young children with CP is often passive in nature, typically waiting until clinical signs appear, then focussing on static stretching, the traditional Neural Developmental Therapy (NDT), bracing with ankle-foot orthoses and botulinum toxin injection of spastic muscles^13–15^, a “wait-and-see”approach^16^. Yet targeted, walking training in school-aged children improves walking performance in children with CP^17,18^. The passive, infrequent and delayed approach to treatment of the lower extremity for young children with hemiparetic CP is in contrast to evidence from animal models of early brain injury, which demonstrates the importance of early, intensive rehabilitation.

Inactivating the primary motor cortex in kittens during a critical period of development impairs the development of motor circuits and motor function^19,20^. Initiating motor training of the affected limb while kittens are young improves motor function and the integrity of motor circuits, whereas training at an older age is less effective^21^. Critical periods of lower extremity motor development in the human may occur before the age of two years, because post-mortem studies have shown mature myelin on the corticospinal tract at the lumbar neurological level around this age^22^. Since mature myelin is associated with reduced neuroplasticity^23^, we suggest that plasticity would be greatest prior to the emergence of mature myelin. Therapeutic approaches to enhance developmental neuroplasticity of children with perinatal stroke have been reviewed recently^24^. In a pilot study, we showed that intensive activity-based rehabilitation for the lower extremity in children with hemiparesis under the age of two resulted in large improvements in walking^25^.

We hypothesized that early, intensive, child-initiated therapy for the lower extremity in children with perinatal stroke would result in greater improvements in motor function than usual care. In this paper, we focus on the changes in gross motor function.

## Methods

The protocol of the study has been described^26^ and will be recounted briefly here. The study was approved by the Health Research Ethics Board at the University of Alberta (Pro00032297).

Written consent was provided by parents/guardians of all participants.

### Participants

#### Inclusion criteria

1. Hemiparesis with MRI-confirmed perinatal stroke, categorized as neonatal arterial ischemic stroke (NAIS), arterial presumed perinatal ischemic stroke (APPIS), or periventricular venous infarct (PVI)^27^.
2. Born at ≥32 weeks gestation, with age at entry to study between 8 months and 3.0 years old.
3. No other neurological disorders.
4. Parent/guardian able to attend all tests and training.
5. Written, informed consent from parent/guardian.

We focus on ischemic, perinatal stroke to reduce confounds associated with other causes of hemiparetic CP. The lower age limit was based on our estimate that an 8-month-old would be able to participate in the intervention for an hour, and the upper limit was to include children above our estimated critical period to determine if age is a factor.

#### Exclusion criteria

1. Extensive brain injuries beyond unilateral ischemic stroke.
2. Musculoskeletal, cognitive or behavioral impairments that preclude participation in the protocol.
3. Unstable epileptic seizures within the past 6 months or taking anti-epileptic medication.
4. Any contraindications to transcranial magnetic stimulation (TMS), because TMS was an outcome measure.
5. Botulinum toxin injection or surgery in the lower extremity in the past 6 months.

Clinical partners identified participants and potentially suitable participants were screened in person by a research physical therapist (PT). Evidence of perinatal stroke on MRI was confirmed by a pediatric neurologist (AK).

### Trial design

A waitlist control, single blind, randomized controlled trial (RCT) was conducted at two centres (Edmonton and Calgary, Alberta, Canada). The experimental design is represented in Figure 1A. Children were stratified by city, then randomized to train immediately (Immediate Group) or delay training for 3 months (Delay Group). Children in an additional parallel cohort who lived beyond commuting distance were trained immediately by their parent or guardian (Parent-trained Group) with guidance from a PT. Final follow-up occurred within three months of each child’s fourth birthday.

**Figure 1.**
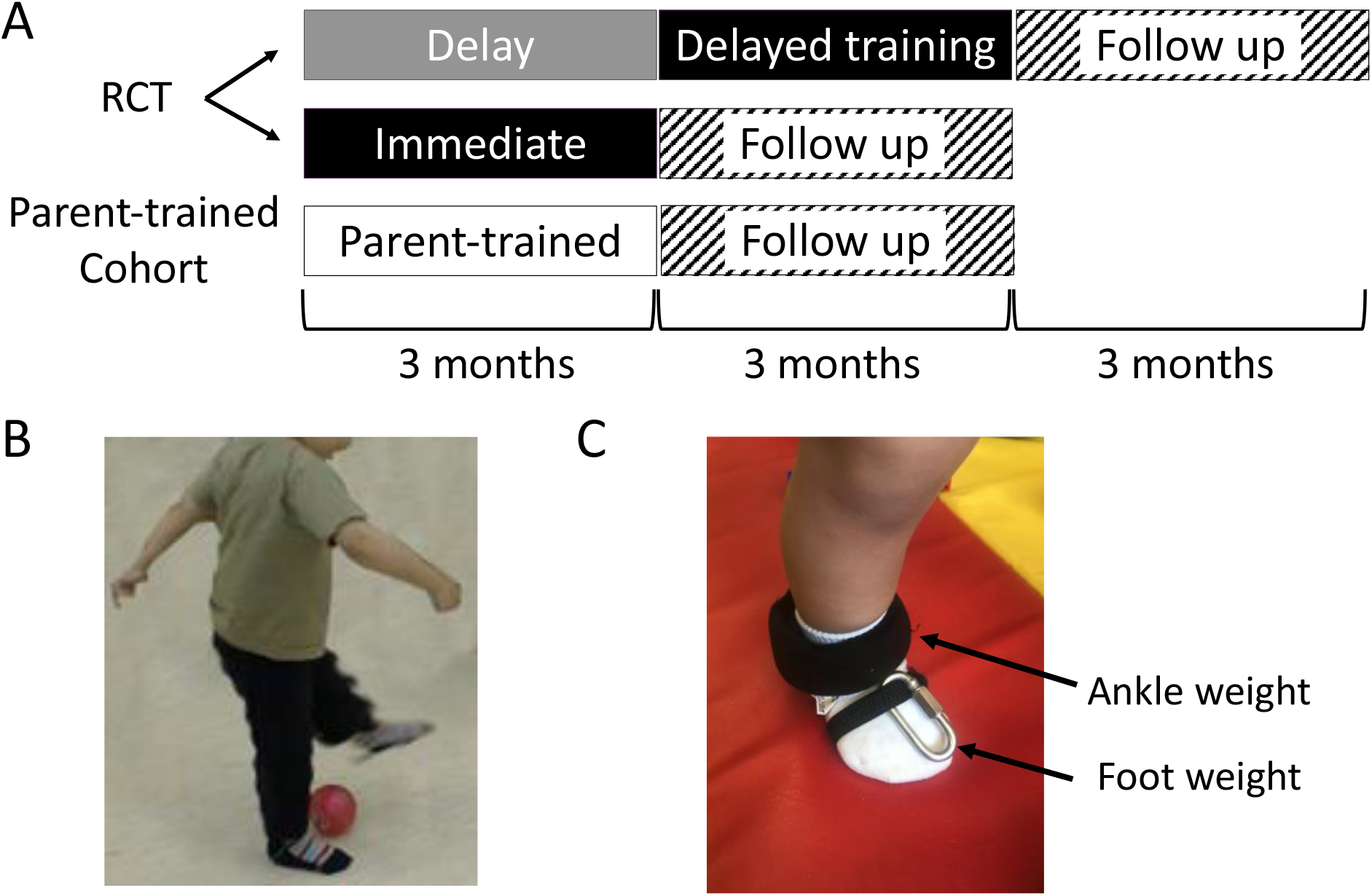
Experimental Design and Methods. A) A schematic diagram of the experimental design indicating the RCT component (top two rows) and the Parent-trained Group (third row). B) A child demonstrating an example of an activity performed during training, reproduced with permission from Oxford University Press^26^. C) Ankle weights were fastened around the lower leg for added strength training, and 1/4” chain links were used on the foot to strengthen dorsiflexors. Abbreviations: RCT, randomized controlled trial.

### Randomization

Participant flow and sample size are shown in a CONSORT diagram (Figure 2). Participants completed all baseline testing before being randomized into either Immediate or Delayed-training Groups in a 1:1 allocation ratio. A biostatistician generated the group allocation sequence using a computer-generated permuted block design with a block size of 2 to 8. Group assignment was concealed in sequentially numbered, sealed envelopes.

**Figure 2.**
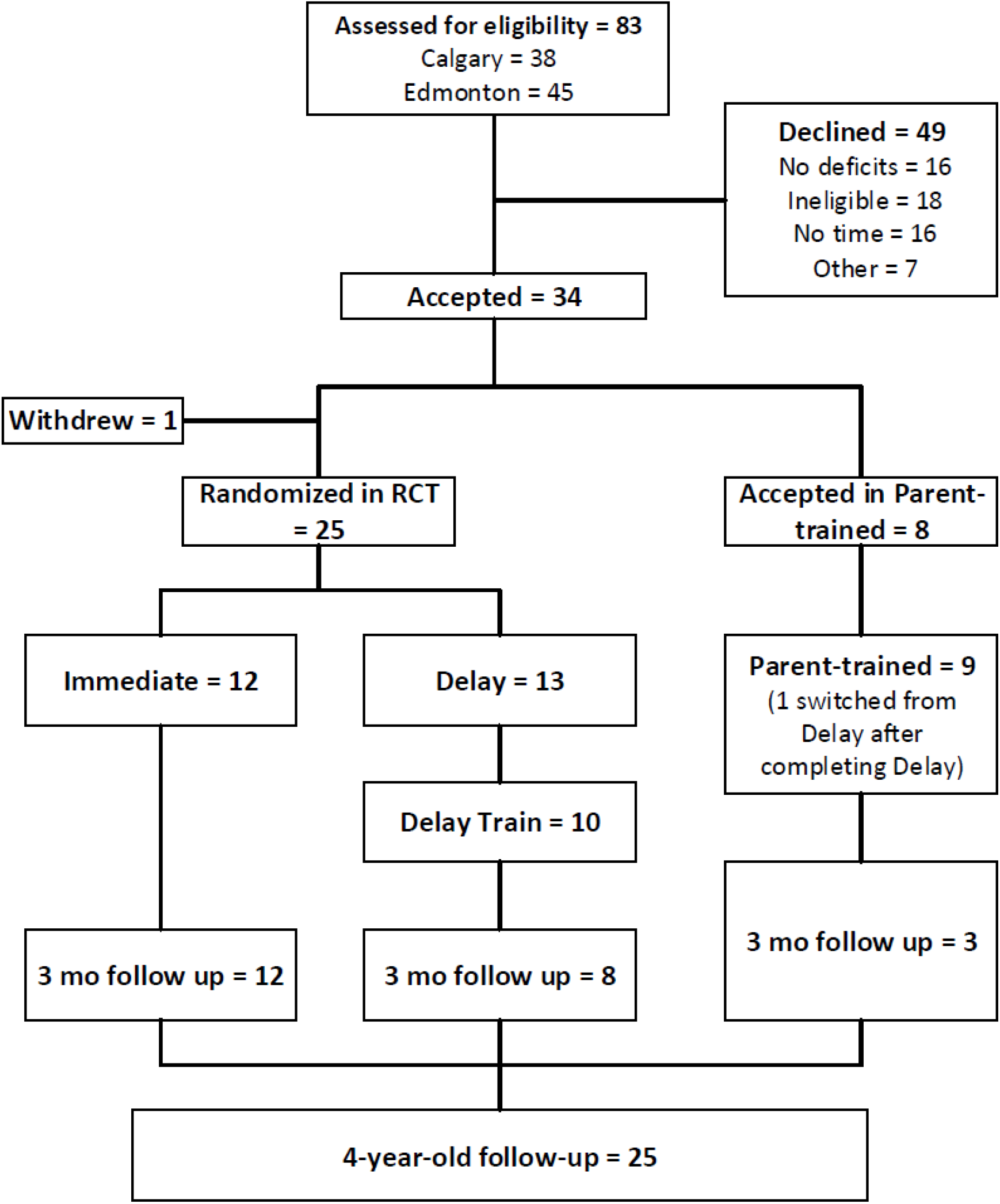
CONSORT Flow Diagram. Eligible children entered the randomized controlled trial (RCT) or the Parent-trained cohort if they live beyond commuting distance. Children in the RCT were allocated to either train immediately (Immediate Group) or delay training for 3 months (Delay Group). The Delay Group served as a usual care control and were trained after the delay period. Children were followed for 3 months after training and reassessed at 4 years old.

### Intervention

The ELEVATE (Engaging the Lower Extremity Via Active Therapy Early) intervention was 1 hour/day, 4 days/week for 12 weeks. The intervention was delivered by one of two PTs (DL or KB) in a laboratory at the University of Alberta (Edmonton) or a satellite campus in Calgary. In the Parent-trained cohort, the intervention was delivered by the child’s parent in their home, guided by one of the two therapists, depending on the proximity of the child’s home to each centre.

The intervention was intensive, child-initiated, play-based movement of the lower extremities, as described in the protocol publication^26^. The intervention occurred primarily over ground on various surfaces, including linoleum, carpet, mats, grass, and cement sidewalks.

Children occasionally walked on a treadmill for parts of the training sessions if that was their preference. Children wore soft-soled slippers and no orthotics while training to enhance the use of muscles in the feet and around the ankle. Activities included ascending and descending stairs and ramps, walking on stable and unstable surfaces, stepping over obstacles, balancing in standing, kicking, and squatting to pick up items (Figure 1B). Manual support from the therapist was provided as needed to facilitate 60 minutes of child-initiated activity. To increase exercise intensity and to augment movement error^28^, weights were placed on the dorsum of the foot and the ankle of the affected leg of children with sufficient endurance to stay active for 50 to 60 minutes. Commercially available weights in increments of 110 g were used for the ankle, and one-quarter-inch chain links, ∼20 g each, were affixed to the dorsum of the foot with elasticized fabric (Figure 1C). Parents of the Parent-trained children were provided with the weights and the training therapist contacted them weekly by phone to provide support. In addition, a monthly in-person collaborative training session between the therapist and parent was provided when children returned to the laboratory for assessments. If parents requested more frequent in-person training, it was accommodated.

### Outcomes

#### GMFM

The primary outcome measure was the Gross Motor Function Measure-66 (GMFM-66)^29^. GMFM-66 is a criterion-referenced observational measure to assess change in gross motor function in children with CP. Sixty-six tasks spanning five dimensions were scored, including lying and rolling, sitting, crawling and kneeling, standing, and walking/running/jumping. Reliability, validity, and responsiveness to change have been established for children ≥6 months of age^30,31^. GMFM-66 assessments were performed by pediatric PTs who were blinded to the child’s group assignment. GMFM-66 assessments were performed twice at baseline, separated by a week and monthly thereafter (throughout the study period, see Figure 1A). The two baseline assessments were averaged for each participant.

#### Step Counts

Step counts were assessed using the StepWatch activity monitor (Orthocare Innovations, Seattle, WA). Full-day step counts were recorded at the beginning, middle and end of each study period (delay, training and follow-up) as a measure of activity outside training sessions. Parents placed step counters on the ankle of the child’s affected leg for waking hours (excluding water activities) for a target of six days and documented activities and time of day in a log. Counts from a minimum of two full days of recording were averaged. Step counts were also measured using the StepWatch during every training session to quantify the amount of activity on the affected lower extremity for both therapist-trained and parent-trained children.

#### Kinematics of Walking

Secondary outcomes included assessment of gait while walking on a treadmill. Experimenters for these outcomes were not blinded. Children walked with soft-soled slippers, played with toys on a table in front of them and received manual support on their lateral thorax, as needed. Gait assessments were performed once at baseline and monthly thereafter throughout the study.

A custom-built split-belt treadmill, with a force plate under each belt, was used in Edmonton. A commercial treadmill (TR1200B; LifeSpan, Salt Lake City, UT) without force plates was used in Calgary. Walking kinematics were captured either with the 3-D Investigator (NDI, Waterloo, Ontario, Canada) in Edmonton or the Motion Analysis System using 12 Eagle-4 cameras (Santa Rosa, CA, USA) in Calgary. Markers were placed at the top of the iliac crest, greater trochanter, knee joint line, lateral malleolus, and head of the 5^th^ metatarsal, bilaterally. Customized MatLab script calculated toe clearance (maximum vertical height of the toe marker during the swing phase) and weight-bearing (mean vertical force during stance phase).

In each assessment, children walked for two trials of approximately one minute each at three different speeds, for a total of 6 trials per assessment. We focused on the speed that included at least 10 steps at all assessments throughout the study, typically the median speed of 0.4m/s or 0.6m/s.

We quantified the symmetry of toe clearance using the following formula:

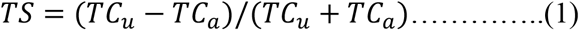

where *TS* is symmetry of toe clearance, *TC*_*u*_ and *TC*_*a*_ are the vertical toe clearance of the unaffected and affected foot, respectively. The percent of a child’s weight borne by the lower extremities in walking was calculated as:

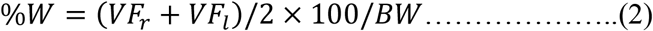

where %*W* is the percent of body weight supported on their feet, *VF*_*r*_ and *VF*_*l*_ are the average vertical forces during the stance phase from the right and left force plates, respectively, and BW is total body weight as measured on a scale.

### Sample size

Pilot data from 4 children suggested that the effect size of the primary outcome measure (GMFM-66) was 1.1^25^. This was based on a GMFM-66 change of 5.5 points, and the predicted change over 3 months without intervention of approximately 3.5, for children with a Gross Motor Function Classification System (GMFCS) level I or II at approximately 1.5 years old. The standard deviation of the change score was estimated to be 2, resulting in a sample size of approximately 16 per group. Formal stopping guidelines were not established because this study evaluated an intervention that did not present increased risk to participants beyond usual care. Clinical decision-making was used to screen for potential overuse injuries and prevent injurious falls throughout training.

### Blinding

Assessors of the primary outcome measure, GMFM-66, were blinded to group allocation but it was not possible to blind treating therapists or parents of participants to group allocation.

### Statistical Methods

Statistical analyses for primary and secondary outcome measures were performed using SAS Ver. 9.4 (SAS Institute Inc., Cary, NC). A multivariable linear regression model was used to examine the effect of age and group allocation on change in GMFM score and toe clearance from baseline to 3 months. All other analyses were conducted using SPSS 20.0 software, including descriptive statistics (mean, standard deviation [SD], and standard error of the mean [SEM]). For participant characteristics, one-way ANOVAs were used to analyze group differences for continuous variables, whereas chi squared tests were used to analyze group differences for categorical data.

## Results

### Recruitment

Recruitment began in 2012 and ended in March, 2018. The recruitment ended at the end of the funding period, at which point we were approaching but below the targeted sample size. The final three month follow-up assessments occurred in July 2019. The final four-year-old follow-up assessment was completed in June 2021.

### Participant flow

The CONSORT diagram (Figure 2), shows 83 potential participants were referred to the study (45 in Edmonton, 38 in Calgary) and screened by a research PT. A total of 24 did not meet inclusion criteria, the most common reasons being no deficits observed (n=14), other neurological complications (n=3) or not ischemic perinatal stroke (n=3). A further 25 potential participants declined to enroll in the study, with the most common reason reported by parents being a prohibitive time commitment (n=16). Thirty-four children (21 in Edmonton, 13 in Calgary) were accepted into the study. One participant in the Immediate Group withdrew before the intervention began due to the child’s unwillingness to participate in assessments. This participant was excluded from the analysis because no data were collected. The total number of participants included in the Delay, Immediate and Parent-trained Groups in the first 3-month period was 13, 12 and 9, respectively. One child withdrew following the three-month delay because the family moved out of the province. One child in the Immediate Group missed the assessments at the end of training due to a family issue unrelated to the study so their data were not included in the statistical analysis, but the child returned for three-month follow-up assessments. Ten children in the Delay Group participated in training following the delay period. One child switched from the Delay Group to the Parent-trained Group after completing the delay period because the family moved out of town. All participants in the Immediate Group completed monthly follow-up for three months following training. The study design did not initially include three-month follow-up for the Delay and Parent-trained Groups but was amended midway through to monitor their function following the training. Hence, three-month follow-up was conducted for only 8 participants in the Delay Group and 3 participants in the Parent-trained Group. Twenty-five children completed a 4-year-old follow-up GMFM-66 assessment. The most common reason for loss to follow-up was scheduling difficulties. Other reasons included refusal to subject child to further assessments, moving out of town following the study and inability to collect data due to COVID-19 restrictions.

### Baseline data

Baseline demographics are shown in Table 1. The average age (months) at baseline was significantly different between the groups (p=0.03), whereas the proportion of females was not significantly (Delay=0.31, Immediate=0.42 and Parent-trained=0.56; p=0.51). The proportion of children classified as Gross Motor Function Classification System (GMFCS) level I versus II when they entered the study was not significantly different between groups (Delay=0.85, Immediate=1.00 and Parent-trained=0.78; p=0.26). All children were classified as GMFCS level I at their four year old follow-up.

**Table 1.**
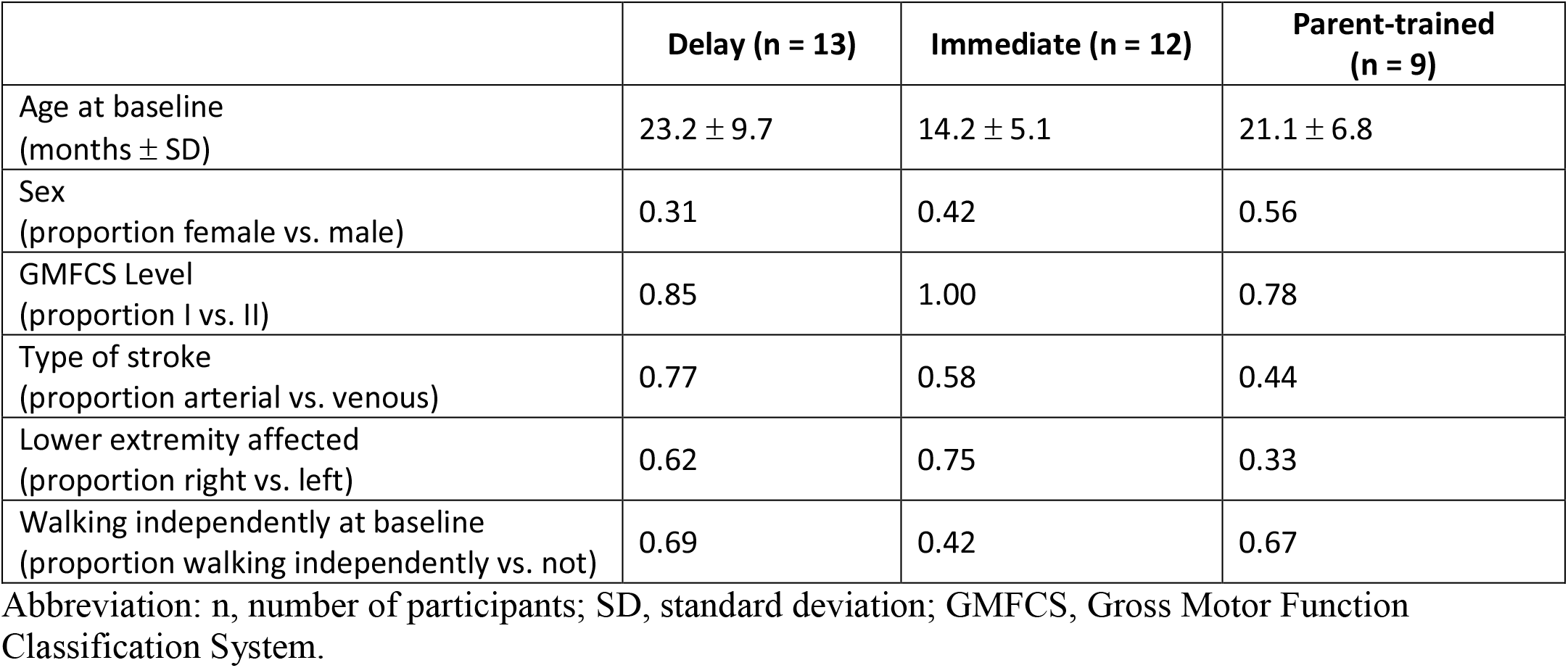
Participant demographics

### Gross Motor Function Measure-66 (GMFM-66)

The average change in GMFM-66 score over the first 3 months was 2.69±2.65 for the Delay Group, 6.62±2.27, for the Immediate Group, and 4.58±3.02 for the Parent-trained Group (Figure 3A). Since there was a difference in age between the Delay and Immediate Groups, GMFM-66 results were analyzed in a multivariable model with both age and group as independent variables. Group allocation but not age was statistically significant. Children in the Immediate Group on average had a change in GMFM score 3.4 points higher (95% confidence interval 1.08-5.64, p=0.006) than children in the Delay Group, adjusted for age. The change in GMFM-66 scores over time in the study for the Delay and Immediate Groups is shown in Figure 3B. Because of the low numbers of participants in each group, no additional statistical comparisons were made. Nevertheless, the descriptive data suggests a continued improvement in the Immediate Group over the follow-up period, while the Delay Group showed smaller improvements in the same period. Further, the changes (Δ) in standing and walking subscores mirrored the change in total GMFM-66 scores (standing: ΔDelay=7.20±7.98, ΔImmediate=24.24±15.88, ΔParent=16.97±18.24; walking: ΔDelay=6.38±8.86, ΔImmediate=11.17±6.18, ΔParent=8.49±6.96).

**Figure 3.**
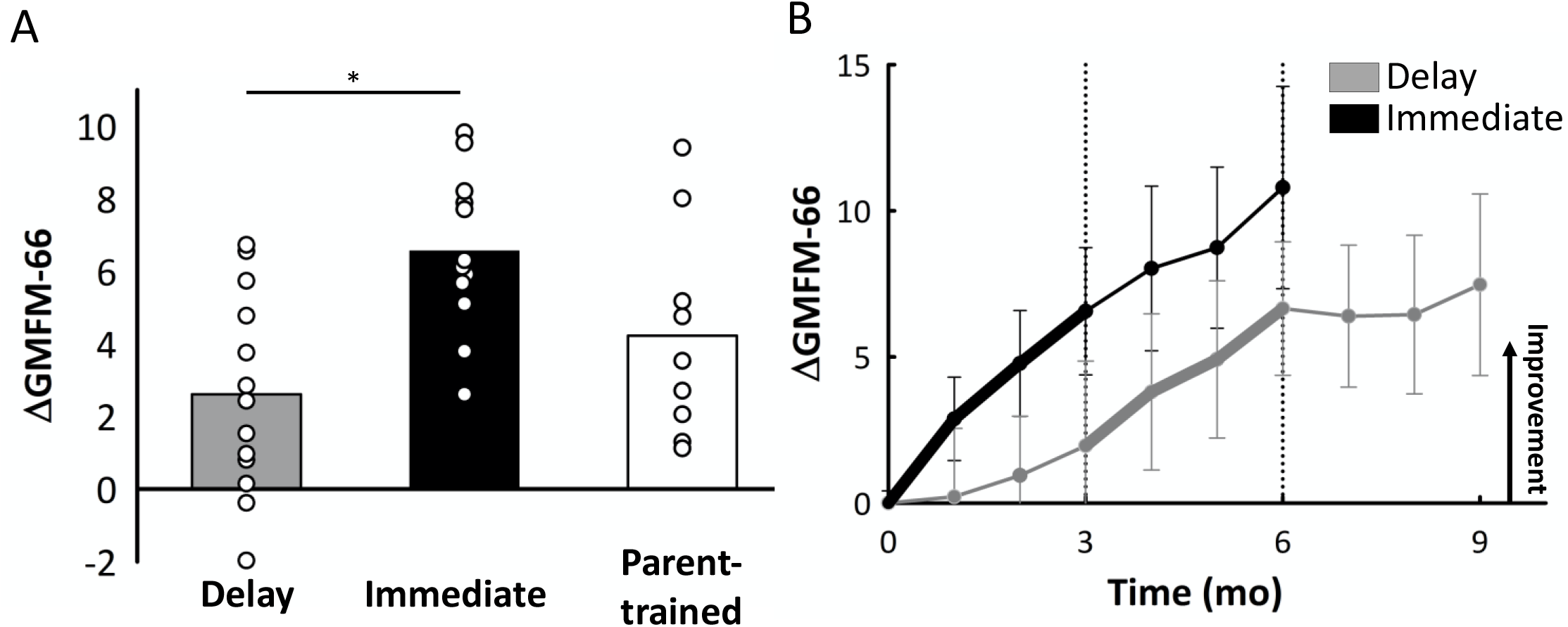
Primary outcome measure: change in Gross Motor Function Measure-66 (GMFM-66) A) Average change scores (i.e., difference between measures at three months and baseline) for the total GMFM-66 score are shown by the bars for each group and change scores for each participant are shown by the white circles. Positive values indicate improvement. Asterisk indicates p<0.05. B) Average change scores for the total GMFM-66 score are indicated for each month for the Delay Group (grey; n=8 for 3 month follow-up) and Immediate Group (black; n=12 for 3 month follow-up) for every month. Bold lines represent the three-month training period, error bars represent standard deviation. Vertical dotted lines segment the 3-month time periods in the study. Abbreviations: GMFM-66, Gross Motor Function Measure-66

At the four-year-old follow-up, the average GMFM-66 score was 69.6±5.8 for children who had been in the Delay Group (n=9), 74.5±7.6 for those in the Immediate Group (n=9) and 73.3±6.5 for the Parent-trained Group (n=7). These mean scores fall between the 40^th^ and 65^th^ percentiles for children classified as GMFCS level I at four years old in the reference curves reported by Hanna and colleagues^32^. The percentile scores for most children at the beginning of the study are unknown because there are no percentile scores for children younger than 2 years old.

### Step Counts

Step counts were used as a measure of training intensity. Average step counts on the affected lower extremity during the first and last week of training are illustrated in Figure 4. The average number of steps increased over the training period for all children in the RCT who were trained by a PT. The average step counts in the Parent-trained Group, however, increased over the training period for children who were not walking independently at the beginning of training but did not increase among children who were walking independently (details in Supplementary material, Table 1). The average age of independent walking, defined as the ability to take 10 steps forward without assistance, assessed with the GMFM-66 or reported by their parents, was 19.8±6.3, 15.3±2.5 and 19.0±4.3 (mean months ± SD) for the Delay, Immediate and Parent-trained Groups, respectively.

**Figure 4.**
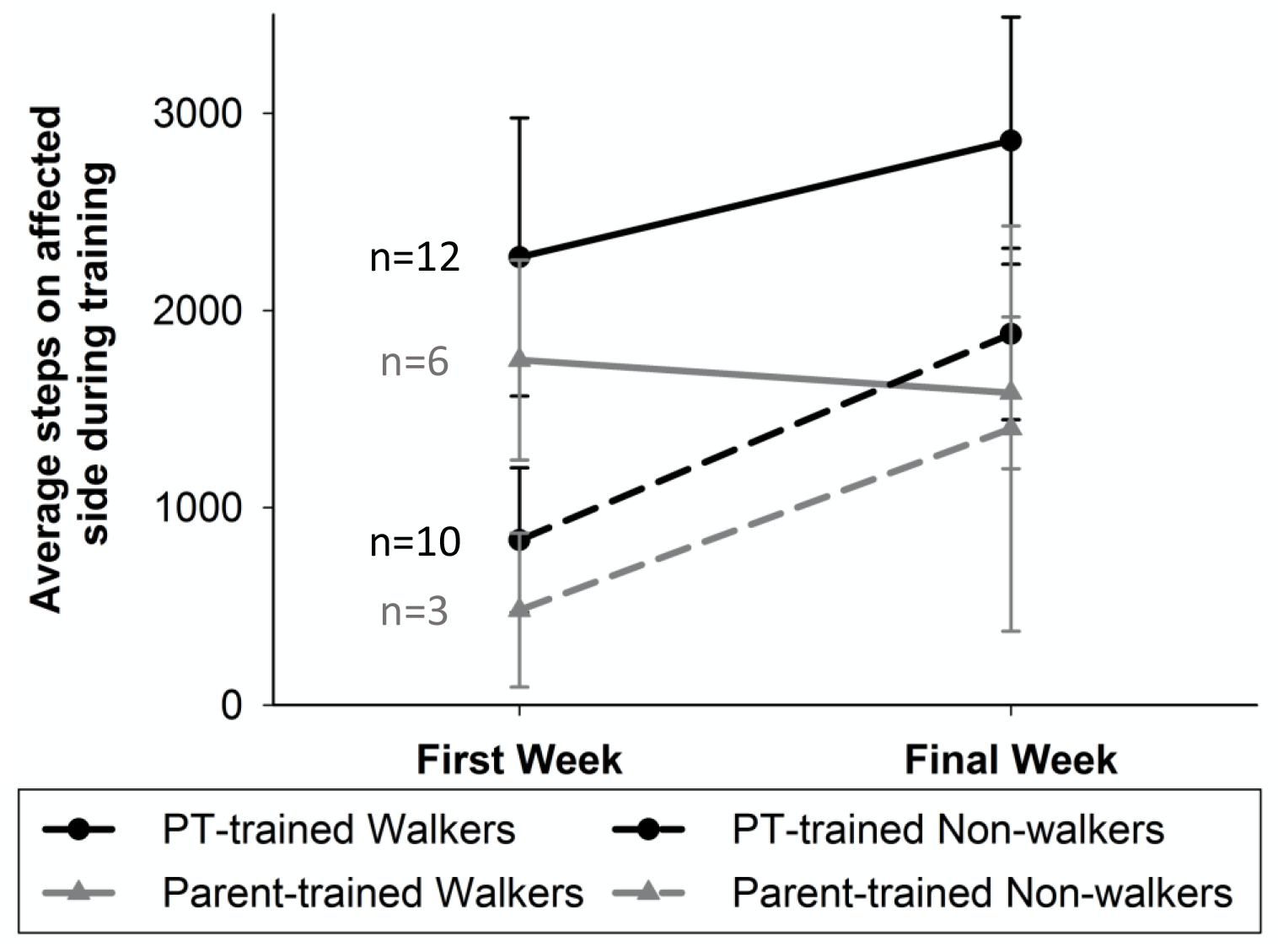
Step counts during training. Black circles represent the average step count on the affected lower extremity for PT-trained children (including both immediately and delay-trained) during the first and final week of training. Gray triangles represent the average step counts for children in the Parent-trained Group. Solid lines represent children who were able to walk independently during the first week of training and dashed lines indicate children who required assistance to walk at the beginning of training. Error bars represent standard deviation. Abbreviations: PT, physical therapist.

Full-day step counts were used to gauge the level of activity in children on non-training days. In general, children with perinatal stroke showed increases of about 1000 steps/day over a 3-month period with or without training, with considerable variation between children (Supplementary material, Figure 1).

### Kinematics of Walking

Perfect symmetry in toe clearance of the left and right foot is represented by zero (Equation 1). Since asymmetry could be positive or negative depending on whether the unaffected or affected toe is lifted higher, the change in the symmetry of toe clearance (ΔTS) over three months was calculated as: *ΔTS* = |*TS*_*b*_| − |*TS*_*e*_|, where *TS*_*b*_ is the symmetry at baseline, and *TS*_*e*_ is the symmetry at the end of the three-month period (i.e. delay or training). Thus, a positive *ΔTS* means they became more symmetric, and a negative *ΔTS* means they became more asymmetric. The average change in symmetry of toe clearance (Figure 5A) worsened for the Delay Group (−0.08±0.13), improved for the Immediate Group (0.07±0.12) and changed minimally for the Parent-trained Group (−0.01±0.10). Since there was a significant difference in age between the Delay and Immediate groups, statistical analysis was performed using a multivariable model. In the multivariable model with both age and group as independent variables, group was not statistically significant (p=0.10).

**Figure 5.**
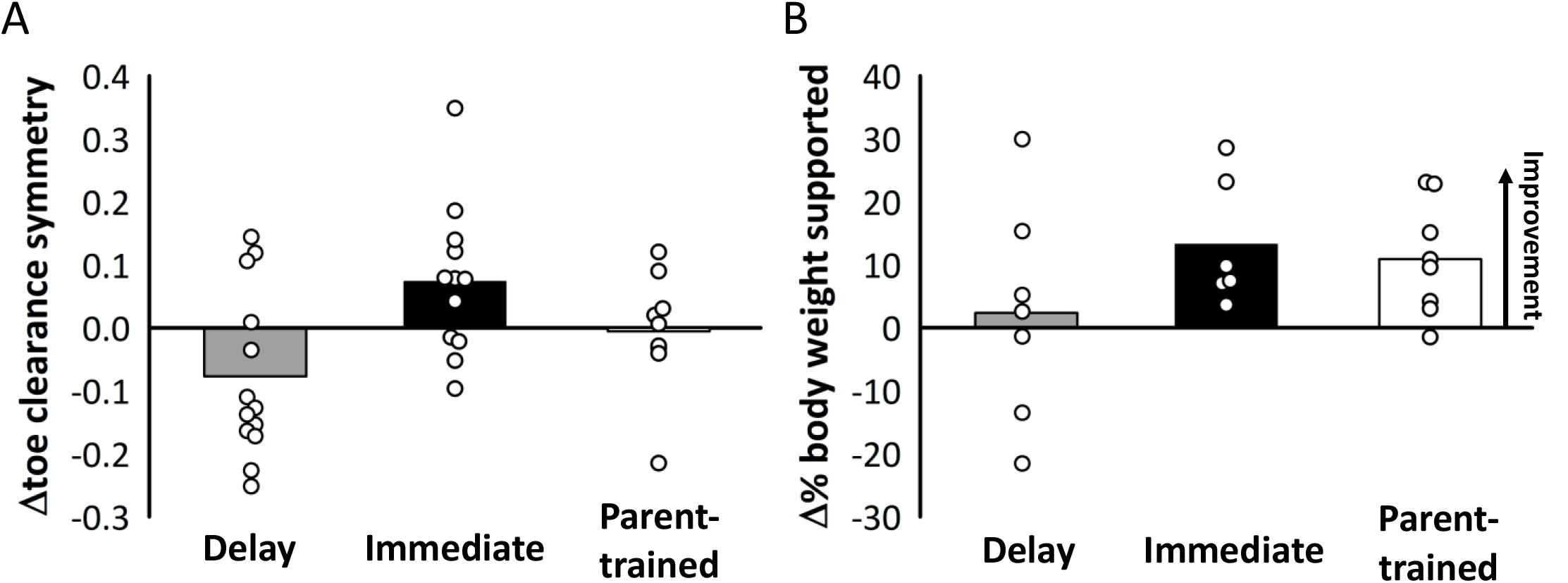
Treadmill Walking. A) Average change scores (i.e., difference between measures at three months and baseline) for the toe clearance symmetry score are shown by the bars for each group and change scores for each participant are shown by the white circles. Delay n=13, Immediate n=12, Parent-trained n=9. B) Average change scores for the percent of their body weight a child could independently support are shown by the bars for each group and change scores for each participant are shown by the white circles. Positive values indicate improvement in both measures. Delay n=7, Immediate n=6, Parent-trained n=8.

Weight-bearing during walking was expressed as a percentage of the child’s body weight. Independent weight-bearing will approach or exceed 100%, so increases in weight-bearing indicated more proficient walkers. Weight-bearing increased by 2.4%±7.2 for children in the Delay Group, 13.3%±10.1 for the Immediate Group and 10.9%±9.0 for children in the Parent-trained Group (Figure 5B). Only descriptive statistics were performed since a treadmill with force plates was only available to children undergoing assessments in Edmonton (n=21).

### Harms

No unintended effects were observed in any participants.

## Discussion

The intensive ELEVATE rehabilitation program for young children with perinatal stroke resulted in significant improvement in gross motor function as compared to usual care. Children in the Immediate Group on average had a change in GMFM-66 score 3.4 units higher (95% confidence interval 1.08-5.64) than children in the Delay Group adjusted for age. This change is higher than previously reported minimum clinically important differences (MCIDs) of 1.7 (medium) and 2.7 (large)^33^, although these MCIDs were established in an older population of children and included children with unilateral and bilateral CP. By the end of the training, children who were walking independently were taking an average of approximately 3000 steps (2862) with their affected lower extremity in one hour of training, and children who were walking with assistance were taking an average of approximately 2000 steps (1881). There was also a trend for improved symmetry in walking with training, but this was not statistically significant. These results demonstrate that it is possible to engage young children in intensive training of the lower extremity resulting in significant improvement in gross motor function in the short term.

### Limitations

Despite randomization there was a difference in age between the control and intervention groups. Since intervening at an earlier age could be more efficacious, a multivariable statistical modelling was used to account for the age difference. Another limitation is the relatively small number of participants (n=25 in RCT), and equipment limitations that only allowed weight-bearing in walking to be assessed at one experimental site. Further, step counts were used to measure dosage or intensity of the intervention, but the training also included other active while stationary skills such as standing balance and squatting, which are not reflected in step counts. Finally, one 3-month bout of intensive training may not be sufficient to have a significant impact on long-term motor function. Determining the ideal time for additional training is needed.

### Nature of Motor Intervention

The premise of this study is based on enhancing neuroplasticity in the developing nervous system. The development of the nervous system is driven by activity, so a child-initiated, activity-based approach was used. This approach incorporates principles such as specificity, repetition, and intensity to promote experience-dependent plasticity^34^. Novak and colleagues indicated that there is clear evidence for rehabilitation approaches that improve function and those that do not; interventions that use volitional active movements at high intensity to practice task-specific training of real-life activities enhance motor outcomes^35^. Intensive rehabilitation has been shown to result in greater improvements in gross motor function than non-intensive treatment and the effect tends to be stronger for children under two years of age^36^. We observed this trend for larger improvements in GMFM-66 scores when training was initiated before 2 years old (Supplementary Material Figure 2). However, other reasons besides the greater neuroplasticity of a young nervous system should not be discounted, including the natural reduction in improvement of the GMFM-66 with age^32^, the greater challenge in directing children around the age of two years old, and the difficulty of enhancing lower extremity function in independently walking children. Differentiating between these factors is difficult.

ELEVATE was tailored to each child to continually challenge their functional ability. For example, weights were added to the affected foot and ankle and this was increased when the child’s gait became more symmetrical. Improvements in function only continue when difficulty of the training is increased^37^. The challenge in ELEVATE included increasing the number of repetitions (steps) and the presumed activity of the muscles (e.g. use of weights and progressively increasing task difficulty to challenge balance and gross motor skills).

We provided ample opportunities for children to develop motor skills through a variety of sensorimotor experiences, analogous to approaches used by the Neuronal Group Selection Theory^38^. For example, children in our study trained on many walking surfaces. Motivation was increased and reinforced throughout the training using play and positive feedback. Motivation and child-initiated movement are essential to enhance neuroplasticity, so task-specific training that is rewarding and enjoyable may promote higher intensity of training and spontaneous, child-initiated practice of skills outside of training sessions. We speculate that the continued improvement during the follow-up period suggests the children continued to use their newly learned skills. Indeed, the full-day step counts (Supplementary Material, Figure 1) suggest that children in the Immediate and Parent-trained Groups continued to increase in daily steps and GMFM scores (Figure 3B), while the Delay Group showed less improvement in both. The differences may be related to the personal characteristics of the children.

### Early Intervention

The children entered this study as young as eight months old, when typically developing children increase their lower extremity strength as they begin to pull to stand and increase weight-bearing. Lower extremity muscle volume is comparable between typically developing children and those with CP until 15 months of age^39^. The change after 15 months is likely related in part to reduced use as typically developing children begin to walk independently around 12±3 months and involves thousands of steps per day, while those with CP are less active and have fewer opportunities to develop lower extremity strength^40,41^. Although it is unclear whether physical activity prevents contractures, early rehabilitation focusing on using the full range of motion may help, since decreased muscle extensibility has been observed as early as 25 to 30 months of age, with contractures developing before the age of three^39,42^.

Several training-based protocols have been reported or are underway for young children (e.g. GAME, HABIT-ILE, small steps, baby-CIMT, baby-bimanual, reviewed in^15,43^). While most of these early interventions target the upper extremities, some have included the lower extremity^10,11^. Here, we show that early, activity-based intervention focused on the lower extremity can substantially improve gross motor function.

As the efficacy of early interventions is established, early diagnosis of perinatal stroke, and CP in general, is increasingly important (reviewed in^44–46^). Facilitating early detection and diagnosis is especially important for perinatal strokes where approximately half of cases are not identified at the time of birth and children often have few risk factors for CP^47^. The importance of early diagnosis and rehabilitation is reflected in recent recommendations for the diagnosis of CP and referral for early rehabilitation^48^.

### Parent Involvement

Children trained by their parents demonstrated variable consistency and efficacy. Home programs have been recommended to increase hand function and self-care for children with CP^10,35,49^ and are well-received by parents^10^. In addition to increasing the intensity of intervention, home programs offer practice in the natural environment, flexibility with scheduling, and cost-effectiveness^50^. Some of the difficulties parents encountered in the past included compliance pressure and feeling a lack of confidence^51,52^. In our study, it was especially challenging for parents with children already independently walking, as seen from the step counts during training (Figure 4). Training families to administer early interventions at home could empower them with the skills to facilitate their child’s rehabilitation while also enhancing their accessibility to a PT. Parents of two participants in the Parent-trained Group requested closer contact with the PT. This included more frequent visits than the monthly in-person training. Interestingly, the same two children showed the greatest improvement in gross motor function among children in the Parent-trained Group. Thus, more frequent in-person parental support from the therapist may be beneficial.

### Future Directions

An intervention that requires PT training one hour/day, four days per week, is costly and not always feasible for families. Exploring different delivery models, including a hybrid approach of both therapist and parental delivery of intervention may be a good compromise, which we are currently investigating. We are also pursuing the generalizability of ELEVATE for children at risk of bilateral CP from encephalopathy of prematurity.

## Supporting information

CONSORT checklist

ICMJE disclosure form

License to reproduce one figure

## Data Availability

The data will be made available once the manuscript has been accepted for publication by a peer-reviewed journal.

## Acknowledgements

The authors thank Dr. Elizabeth Condliffe for assistance reviewing potential participants for eligibility, Dr. Jerome Yager for assistance with recruitment, Michelle Teves for assistance with data collection and analysis, and all the PT assessors in Edmonton and Calgary. We thank Maryna Yaskina for statistical analysis, which was facilitated by the Women and Children’s Health Research Institute through the generous support of the Stollery Children’s Hospital Foundation. We thank the C.H.Riddell Family Movement Assessment Centre at the Alberta Children’s Hospital for recording the kinematics of treadmill walking in Calgary. We are tremendously grateful to the parents and guardians who dedicated large amounts of time to our study. Without their dedication, this study would not have been possible.

