## Supplementary material for "Early, intensive rehabilitation improves gross motor function after perinatal stroke: results of a randomized controlled trial"

|  | | Physical Therapist-trained | Parent-trained Group |
| --- | --- | --- | --- |
| First week | Independent Walkers | 2271 ± 705 (n=12) | 1749 ± 507 (n=6) |
|  | Non-walkers | 836 ± 367 (n=10) | 481 ± 390 (n=3) |
| Final week | Independent Walkers | 2862 ± 626 (n=12) | 1582 ± 385 (n=6) |
|  | Non-walkers | 1881 ± 434 (n=10) | 1402 ± 1027 (n=3) |

**Supplementary Table 1 – Average step counts on the affected lower extremity during the hour of training (mean ± SD).** Physical Therapist-trained include children from both the Immediate and Delay Groups, during their respective training periods.

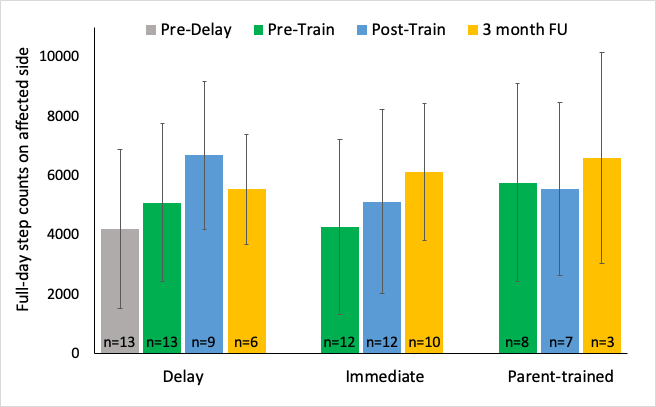

**Supplementary Figure 1 – Full-day step counts on the affected lower extremity.** Bars indicate average step counts measured over 2 to 6 full days for each child then averaged across children in the same group for each phase of the study. The number of children from which we obtained valid measures over at least two full days is indicated by n in each bar. Error bars indicate standard deviation.
Abbreviations: FU, follow-up.

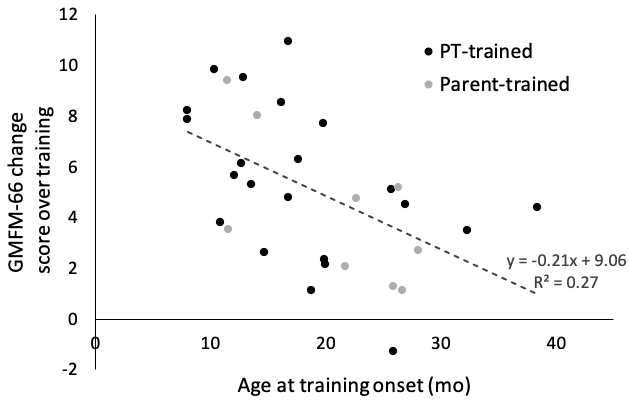

**Supplementary Figure 2 – GMFM-66 change scores over three months of training versus age (months) at the onset of training.** Black markers indicate change scores for children trained by a PT (n=22), i.e., both from the Delay and Immediate Groups and gray markers indicate changes in children trained by a parent (n=9). The trendline includes all data (n=31; R = -0.52 p=0.003).
