## Supplementary material for "Early, intensive rehabilitation improves gross motor function after perinatal stroke: results of a randomized controlled trial": License to reproduce one figure

OXFORD UNIVERSITY PRESS LICENSE
TERMS AND CONDITIONS

Jul 21, 2021

---

This Agreement between Caitlin Hurd ("You") and Oxford University Press ("Oxford University Press") consists of your license details and the terms and conditions provided by Oxford University Press and Copyright Clearance Center.

| | |
|---|---|
| License Number | 5113710177157 |
| License date | Jul 21, 2021 |
| Licensed Content Publisher | Oxford University Press |
| Licensed Content Publication | Physical Therapy |
| Licensed Content Title | Early Intensive Leg Training to Enhance Walking in Children With Perinatal Stroke: Protocol for a Randomized Controlled Trial |
| Licensed Content Author | Hurd, Caitlin; Livingstone, Donna |
| Licensed Content Date | May 8, 2017 |

| | |
|---|---|
| Licensed Content Volume | 97 |
| Licensed Content Issue | 8 |
| Type of Use | Journal |
| Requestor type | Author of this OUP content |
| Pharmaceutical support or sponsorship for this project | No |
| Format | Electronic |
| Portion | Figure/table |
| Number of figures/tables | 1 |
| Will you be translating? | No |
| Circulation/distribution | 1 |
| Title of new article | Early, intensive rehabilitation improves gross motor function after perinatal stroke: results of a randomized controlled trial |
| Lead author | Caitlin Hurd |
| Title of targeted journal | MedRxiv |

| | |
|---|---|
| Publisher | MedRxiv |
| Expected publication date | Jul 2021 |
| Portions | We are planning to use a single picture from the supplementary file (the first picture on the first page under "kicking"). |
| Requestor Location | Caitlin Hurd<br>3-88 Corbett Hall<br>University of Alberta<br><br>Edmonton, AB T6E 2G4<br>Canada<br>Attn: Caitlin Hurd |
| Publisher Tax ID | GB125506730 |
| Total | 0.00 CAD |

Terms and Conditions

**STANDARD TERMS AND CONDITIONS FOR REPRODUCTION OF MATERIAL FROM AN OXFORD UNIVERSITY PRESS JOURNAL**

1. Use of the material is restricted to the type of use specified in your order details.

2. This permission covers the use of the material in the English language in the following territory: world. If you have requested additional permission to translate this material, the terms and conditions of this reuse will be set out in clause 12.

3. This permission is limited to the particular use authorized in (1) above and does not allow you to sanction its use elsewhere in any other format other than specified above, nor does it

apply to quotations, images, artistic works etc that have been reproduced from other sources which may be part of the material to be used.

4. No alteration, omission or addition is made to the material without our written consent. Permission must be re-cleared with Oxford University Press if/when you decide to reprint.

5. The following credit line appears wherever the material is used: author, title, journal, year, volume, issue number, pagination, by permission of Oxford University Press or the sponsoring society if the journal is a society journal. Where a journal is being published on behalf of a learned society, the details of that society must be included in the credit line.

6. For the reproduction of a full article from an Oxford University Press journal for whatever purpose, the corresponding author of the material concerned should be informed of the proposed use. Contact details for the corresponding authors of all Oxford University Press journal contact can be found alongside either the abstract or full text of the article concerned, accessible from www.oxfordjournals.org Should there be a problem clearing these rights, please contact

7. If the credit line or acknowledgement in our publication indicates that any of the figures, images or photos was reproduced, drawn or modified from an earlier source it will be necessary for you to clear this permission with the original publisher as well. If this permission has not been obtained, please note that this material cannot be included in your publication/photocopies.

8. While you may exercise the rights licensed immediately upon issuance of the license at the end of the licensing process for the transaction, provided that you have disclosed complete and accurate details of your proposed use, no license is finally effective unless and until full payment is received from you (either by Oxford University Press or by Copyright Clearance Center (CCC)) as provided in CCC's Billing and Payment terms and conditions. If full payment is not received on a timely basis, then any license preliminarily granted shall be deemed automatically revoked and shall be void as if never granted. Further, in the event that you breach any of these terms and conditions or any of CCC's Billing and Payment terms and conditions, the license is automatically revoked and shall be void as if never granted. Use of materials as described in a revoked license, as well as any use of the materials beyond the scope of an unrevoked license, may constitute copyright infringement and Oxford University Press reserves the right to take any and all action to protect its copyright in the materials.

9. This license is personal to you and may not be sublicensed, assigned or transferred by you to any other person without Oxford University Press’s written permission.

10. Oxford University Press reserves all rights not specifically granted in the combination of (i) the license details provided by you and accepted in the course of this licensing transaction, (ii) these terms and conditions and (iii) CCC's Billing and Payment terms and conditions.

11. You hereby indemnify and agree to hold harmless Oxford University Press and CCC, and their respective officers, directors, employs and agents, from and against any and all claims arising out of your use of the licensed material other than as specifically authorized pursuant to this license.

12. Other Terms and Conditions:

v1.4

**Questions? or +1-855-239-3415 (toll free in the US) or +1-978-646-2777.**
